# Presence of concurrent sarcoid-like granulomas indicate better survival in cancer patients

**DOI:** 10.1101/2020.03.28.20046003

**Authors:** Mukunthan Murthi, Keiichiro Yoshioka, Jeong Hee Cho, Sixto Arias, Elio Danna, Greg Holt, Koichiro Tatsumi, Takeshi Kawasaki, Mehdi Mirsaeidi

**Author notes:** **Correspondence author:** Dr. Mehdi Mirsaeidi, Division of Pulmonary and Critical Care, University of Miami, Miami, FL, USA.,. **Authors’ contributions:** Mukunthan Murthi, Keiichiro Yoshioka conducted chart review and collected data, data analysis, literature review and helped in manuscript preparation. Jeong Hee Cho provided melanoma dataset and helped to develop first draft of manuscript. Sixto Arias, Elio Danna, Greg Holt reviewed EBUS procedures and helped in analyzing and discussing results and manuscript preparation. Koichiro Tatsumi, and Takeshi Kawasaki assisted in reviewing the subjects’ data accuracy, data analysis, and in manuscript preparation. Mehdi Mirsaeidi conducted literature review, designed the study, conducted exploratory analysis, performed data analysis, and manuscript preparation.

## Abstract

**Introduction:** An increased risk of sarcoidosis and sarcoid-like reactions in subjects with a history of malignancy has been suggested. We assessed the incidence and clinical characteristics of cancer patients with biopsies containing sarcoid-like granulomas on cancer metastasis and patient survival.

**Methods:** This is a retrospective, multicenter, observational study involving patients who underwent Endobronchial Ultrasound (EBUS) at the University of Miami Hospital, Miami Veterans Affairs Medical Center in the USA and Chiba University in Japan. Subjects with a confirmed diagnosis of cancer and who subsequently developed granulomas in different organs were enrolled. The study was registered at Clinicaltrial.gov (NCT03844698).

**Results:** One hundred and thirty-three patients met the study’s criteria. The most common primary cancer sites were the skin (22.5%), breast (20.3%), and lymph node (LN) (12.8%). Twenty-four (18%) patients developed sarcoidal granulomas within 1 year of cancer diagnosis, 54(40.6%) between 1 and 5 years and 49(36.8%) after 5 years. Imaging showed possible sarcoidal granulomas in lymph nodes in 51 cases (38.3%) and lung tissue and mediastinal lymph nodes in 73 cases (54.9%); some parenchymal reticular opacity and fibrosis were found in 5(3.7%) and significant parenchymal fibrosis in 2(1.5%) subjects. According to logistic regression analysis, the frequency of metastatic cancer was significantly lower in patients with sarcoidal granulomas than in controls. Moreover, multivariate Cox proportional hazard analysis showed a significant survival advantage in those with sarcoidal granuloma.

**Conclusion:** Sarcoidal granulomas are uncommon pathology findings in cancer patients. There is a significant association between the presence of granulomas and reduced metastasis and increased survival. Further study is warranted to understand the protective mechanism involved.

**Take home message:** Our findings suggest that patients with underlying malignancy who develop sarcoidosis and sarcoid-like reactions have a lower risk of stage 4 metastatic disease and better survival compared to patients who do not develop such granulomatous reactions.

## Introduction

Sarcoidosis is a multisystem granulomatous disease that predominantly affects the lung but can be seen in any organ system[1]. The fundamental abnormality in sarcoidosis is the development of pathological structures known as non-caseating granulomas [2] consisting of collections of macrophages and epithelioid cells surrounded by lymphocytes and fibroblasts. Although the etiology of such granuloma formation in sarcoidosis remains unclear, it is believed to be due to an abnormal host immune response to an unknown antigen in genetically susceptible individuals.

Multiple studies have demonstrated an increased risk of cancer following sarcoidosis. In a retrospective study of nearly 9000 sarcoidosis patients, Askling *et al*. reported an elevated risk of cancer with a standardized incidence of 1.3 (CI-1.2-1.4)[3]. In a recent meta-analysis of over 25,000 patients, there was a relative risk of 1.19 (95% CI, 1.07-1.32) for the development of all types of invasive cancers in patients with sarcoidosis[4].

Less is known about the development of sarcoidosis after the onset of malignancy. Patients with an underlying malignancy develop noncaseating epithelioid cell granulomas in regional and distant lymph nodes and/or parenchyma of various organs [5]. Given the characteristic enlargement of lymph nodes associated with sarcoidosis, the clinical and radiological features of these conditions mimic metastatic cancer and require biopsy. To date, only a few studies have analyzed the occurrence of sarcoidosis after cancer diagnosis and most are case reports. We previously described the increased incidence of sarcoidosis among subjects with breast cancer in our registry[6]. Whether the presence of sarcoid-like granulomas affects survival if developed after cancer onset is difficult to assess as different studies have reported conflicting results[7–11].

This study aimed to compare the clinicopathological characteristics and outcomes in cancer patients with and without sarcoid-like granulomas in biopsy specimens. sarcoid-like granulomas are thought to develop from a persistent immune reaction, we postulated that cancer patient’s granulomatous reactions indicate a robust immune response and may provide a survival advantage.

## Methods

### Study Design

We conducted a multicenter, retrospective observational study of over 1600 subjects at the University of Miami Hospital, Miami Veterans Affairs Medical Center (VAMC) in Florida, the United States, and Chiba University in Chiba, Japan. The study was approved by the local Institutional Review Boards (IRBs) with a waiver of informed consent. For better transference, we herein refer to both sarcoidosis and sarcoid reactions occurring in cancer patients as “sarcoidal granuloma”. We identified each case of sarcoidal granuloma by manually sorting through prospectively collected datasets of patients who underwent EBUS-TBNA for suspected cancer diagnosis at the University of Miami, Miami VAMC and Chiba University. A dataset consisting of prospectively collected data for melanoma patients from a single provider at the University of Miami was also used.

### Inclusion and exclusion criteria

Any patient with a pathology finding of sarcoidal granuloma during or after cancer diagnosis was included in the study. Subjects found to have sarcoidal granuloma before the diagnosis of cancer were excluded. Other causes of granulomatous disease were excluded by mycobacterial and fungal staining of pathology samples and culture, review of the subject history and follow-up. For each patient, demographic data, including age, sex and ethnicity, were collected. Details on cancer as well as sarcoidal granulomas were also obtained. All cancers were staged according to the American Joint Committee on Cancer (AJCC) guidelines.

### Survival analysis

Subject group survival was compared to a control group to determine the effect of sarcoidal granulomas on cancer survival. For the ten most commonly occurring cancers among our subjects (breast, bladder, prostate, and pancreatic cancers, lymphoma, RCC, melanoma, colon carcinoma, sarcoma), controls with the same cancer diagnosis were identified and chosen randomly based on the absence of sarcoidal granuloma up to a ratio of 3:1. For both groups, only patients with up-to-date follow-up and for whom their death was documented in the medical records were included for survival analysis. For subjects with more than one cancer diagnosis, cancer that was diagnosed closest to the time of granuloma diagnosis was used for case and control matching.

### Statistical analysis

Categorical variables are described as counts and percentages; their distribution was compared using odds ratios and tested using the chi-squared test and Cochran-Mantel-Haenszel test or exact tests, as indicated. Continuous variables were analyzed with independent sample t-tests (two-tailed). Cox proportional hazard analysis was performed for all cancers to identify whether potentially confounding variables affect survival independently. The selection of variables was based on a review of the literature with regards to factors affecting survival among cancer patients. For the four common cancers among our subjects, i.e., lung and breast cancers, melanoma and lymphoma, we performed the log-rank test to predict statistically significant differences in survival. Kaplan-Meier curves were generated to illustrate the ordinality of any differences found. We also applied logistic regression analysis to examine the relationship between various clinical factors and the likelihood of stage 4 metastatic disease. All analyses were carried out using SPSS software for Windows, version 26. For all results, significance was considered at p<0.05.

## Results

### Patient characteristics

After reviewing 1546 electronic medical records at the University of Miami, 212 patients had sarcoidal granulomas on pathology reports (Figure 1) and 1219 had a history of cancer. 101 (8.3%) patients developed sarcoidal granulomas after cancer diagnosis. Chiba University contributed 32 subjects for analysis producing a total of 133 subjects for the study. Among the 133 subjects, 61 (45.9%) were male and 72 (54.1%) female. The ethnic distribution included 35 (26.3%) Asians, 48 (36.1%) European-Americans, 14 (10.5%) African Americans, 31 (23.3%) Latinos and 5 (3.7%) of mixed ethnicity. The mean age of the subjects was 64.5 years (SD-22 to 86 years). Table 1 summarizes the subjects’ cancer and sarcoidosis characteristics.

**Table 1:**
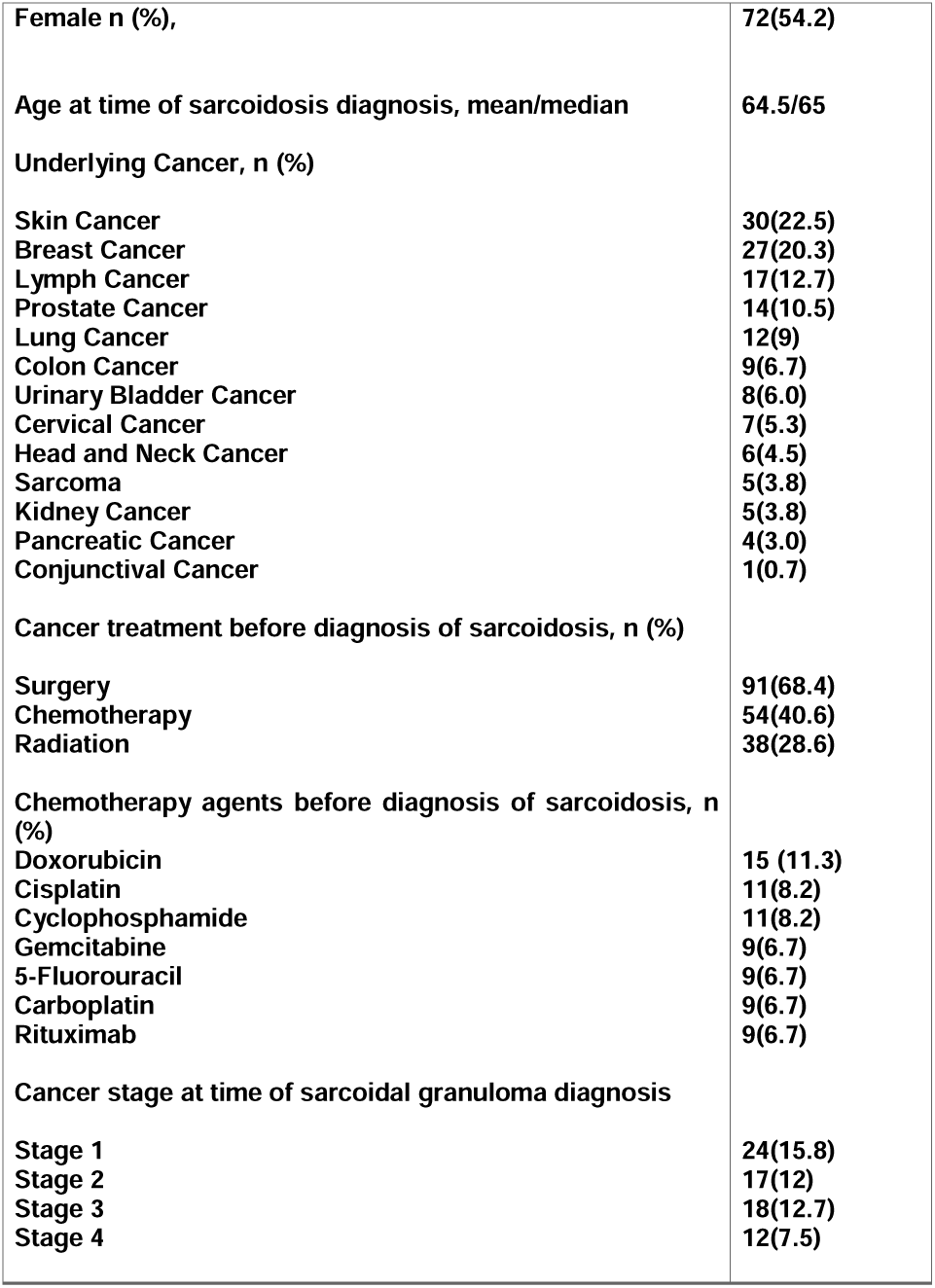
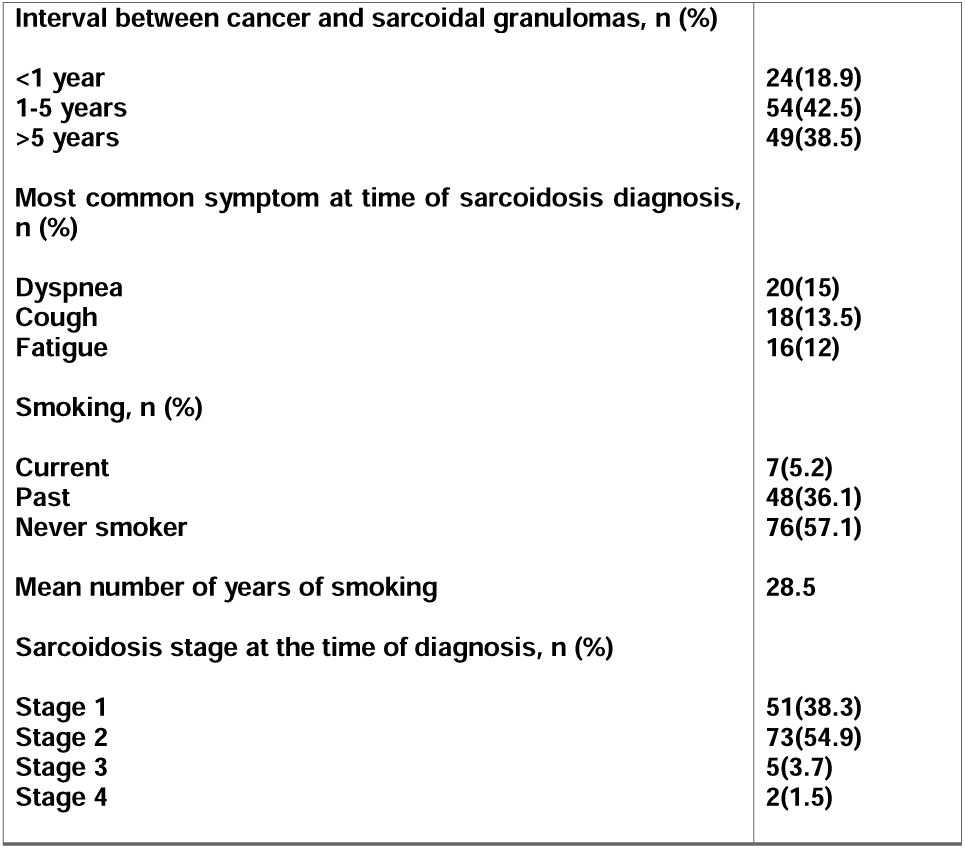
Characteristics of patients with cancer and sarcoidosis.

|  |  |
| --- | --- |
| **Female n (%),** | **72(54.2)** |
| **Age at time of sarcoidosis diagnosis, mean/median** | **64.5/65** |
| **Underlying Cancer, n (%)** |  |
| Skin Cancer | 30(22.5) |
| Breast Cancer | 27(20.3) |
| Lymph Cancer | 17(12.7) |
| Prostate Cancer | 14(10.5) |
| Lung Cancer | 12(9) |
| Colon Cancer | 9(6.7) |
| Urinary Bladder Cancer | 8(6.0) |
| Cervical Cancer | 7(5.3) |
| Head and Neck Cancer | 6(4.5) |
| Sarcoma | 5(3.8) |
| Kidney Cancer | 5(3.8) |
| Pancreatic Cancer | 4(3.0) |
| Conjunctival Cancer | 1(0.7) |
| **Cancer treatment before diagnosis of sarcoidosis, n (%)** |  |
| Surgery | 91(68.4) |
| Chemotherapy | 54(40.6) |
| Radiation | 38(28.6) |
| **Chemotherapy agents before diagnosis of sarcoidosis, n (%)** |  |
| Doxorubicin | 15 (11.3) |
| Cisplatin | 11(8.2) |
| Cyclophosphamide | 11(8.2) |
| Gemcitabine | 9(6.7) |
| 5-Fluorouracil | 9(6.7) |
| Carboplatin | 9(6.7) |
| Rituximab | 9(6.7) |
| **Cancer stage at time of sarcoidal granuloma diagnosis** |  |
| Stage 1 | 24(15.8) |
| Stage 2 | 17(12) |
| Stage 3 | 18(12.7) |
| Stage 4 | 12(7.5) |
| **Interval between cancer and sarcoidal granulomas, n (%)** |  |
| **<1 year** | **24(18.9)** |
| **1-5 years** | **54(42.5)** |
| **>5 years** | **49(38.5)** |
| **Most common symptom at time of sarcoidosis diagnosis, n (%)** |  |
| **Dyspnea** | **20(15)** |
| **Cough** | **18(13.5)** |
| **Fatigue** | **16(12)** |
| **Smoking, n (%)** |  |
| **Current** | **7(5.2)** |
| **Past** | **48(36.1)** |
| **Never smoker** | **76(57.1)** |
| **Mean number of years of smoking** | **28.5** |
| **Sarcoidosis stage at the time of diagnosis, n (%)** |  |
| **Stage 1** | **51(38.3)** |
| **Stage 2** | **73(54.9)** |
| **Stage 3** | **5(3.7)** |
| **Stage 4** | **2(1.5)** |

**Figure 1:**
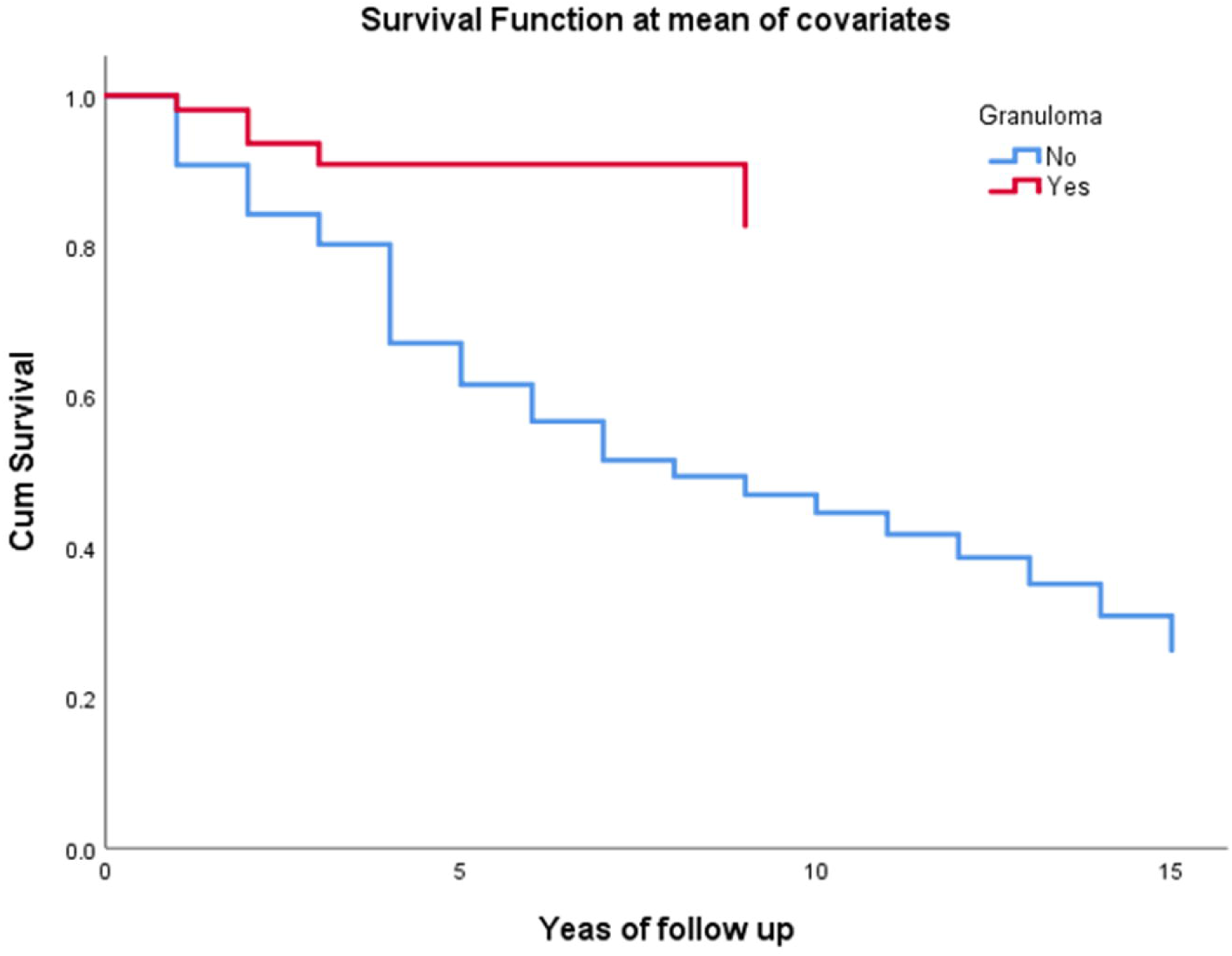
Flowchart showing the selection of cases of cancer with sarcoidal granuloma.

Thirty-nine (29.3%) subjects had a history of more than one cancer before the sarcoidal granuloma diagnosis. The most common primary cancer sites were the skin (30; 22.5%), breast (27; 20.3%), and lymph node (LN) (17; 12.8%). Regarding cancer stages at the time of sarcoidal granuloma diagnosis, 24 cases (15.8%) were stage 1, 17 cases (12%) were stage 2, 18 cases (12.7%) were stage 3 and 12 cases (7.5%) were stage 4.

Fifty-four patients (40.6%) were treated with chemotherapy, 38 patients (28.6%) were treated with radiation and 91 patients (68.4%) underwent surgery prior to sarcoidal granuloma diagnosis. The commonly used chemotherapy agents were doxorubicin (15; 11.3%), cisplatin (11; 8.2%) and cyclophosphamide (11; 8.2%). Other commonly used agents are listed in Table 1. More than half of the subjects (61.4%) developed granulomas within 5 years of their cancer diagnosis. The median interval between cancer and sarcoidosis diagnoses was 3 years (mean 6 years, range 1-37 years).

### Radiological, pathological and laboratory characteristics of subjects with confirmed granulomas

The presence of sarcoidal granulomas in cancer patients led us to define possible sarcoidosis staging by assessing chest imaging via CT or PET/CT. A total of 79% of patients had mediastinal and/or hilar lymph nodes greater than 1 cm at the time of sarcoidal granuloma diagnosis. The commonly enlarged lymph nodes on imaging were station 7 (85 [64%]), 4R (73 [54.9%]), and 2R (50 [37.6%]). The most common parenchymal lesion was the presence of nodules (in 73 subjects; 54.9%), followed by GGO (in 13; 9.7%), reticular opacities (in 9; 6.7%), atelectasis (in 8; 6%) and bronchiectasis (in 3; 2.3%). The mean size of pulmonary nodules was 9.7 mm (range 2-30 mm). Thirty-one (23.3%) patients had extra-thoracic lymph nodes >10 mm or with SUV >2.5. Based on Scadding criteria at the time of diagnosis, if defined them as sarcoidosis, sarcoidosis staging was as follows: stage 1, 38.3%; stage 2, 54.9%; stage 3, 3.7%; and stage 4, 1.5%.

The most common intrathoracic LNs diagnosed with granuloma were station 7 (51.9%), 4R (23.3%), 11L (15.8%) and 11R (15%). Thirty-one (23.3%) subjects had granulomas in the parenchyma. Twenty-six (19.5%) had extra-thoracic granulomas, with skin 10 (7.5%) and liver 5 (3.7%) being the most common sites. Fourteen (10.5%) subjects had necrotizing granulomas on pathology. In total, the frequency of granulomas in our dataset was 8.3%.

Pulmonary function test (PFT) results were available for 61 subjects, 35 (26.3%) of which were normal. In those with impaired function, 12 (9%) had restrictive disease, and 14 (10.5%) had an obstructive pattern. ACE levels were available for 54 patients, and the median value was 25.35 U/L (Mean=31.92). CRP was available for 52 patients, and it was elevated in 21 (15.7%). More than half of our subjects (68; 51.2%) were asymptomatic at the time of sarcoidal granuloma diagnosis. In symptomatic patients, the most common symptoms were dyspnea (in 20; 15%), cough (in 18; 13.5%) and fatigue (in 16; 12%).

### Japanese vs. American patients

We also compared the subject’s nationality to determine whether there is any clinical difference between Japanese or American patients. Japanese subjects were less likely than Americans to have stage 2, 3 and 4 sarcoidosis (OR 0.31 [0.13-0.72], p-value=0.007 and were less likely to develop sarcoidal granulomas within 5 years of cancer diagnosis (OR-0.31 [0.13-0.72] p-value=0.006).

### Association between sarcoidosis granulomas and cancer stage 4 (metastasis)

We compared the characteristics of cancer between the case and control groups. Ten (22.2%) of the subjects in the sarcoidal granuloma group had stage 4 disease compared to 72 (57.6%) subjects in the control group. *Figure 2* depicts the forest plot of the estimates and 95% confidence intervals obtained by logistic regression analysis representing independent factors associated with stage 4 disease in patients with cancer. The presence of granuloma (OR=0.216 [0.097-0.483], p<0.0001) and male gender (0.500 [0.258-0.968], p=0.04) was associated with a lower incidence of stage 4 disease. Of note, there were no significant differences in treatment modality (chemotherapy, radiotherapy, MAB, surgery) between the subjects with and without stage 4 disease. Moreover, we did not find any impact due to the race of the subjects on the cancer stage.

**Figure 2:**
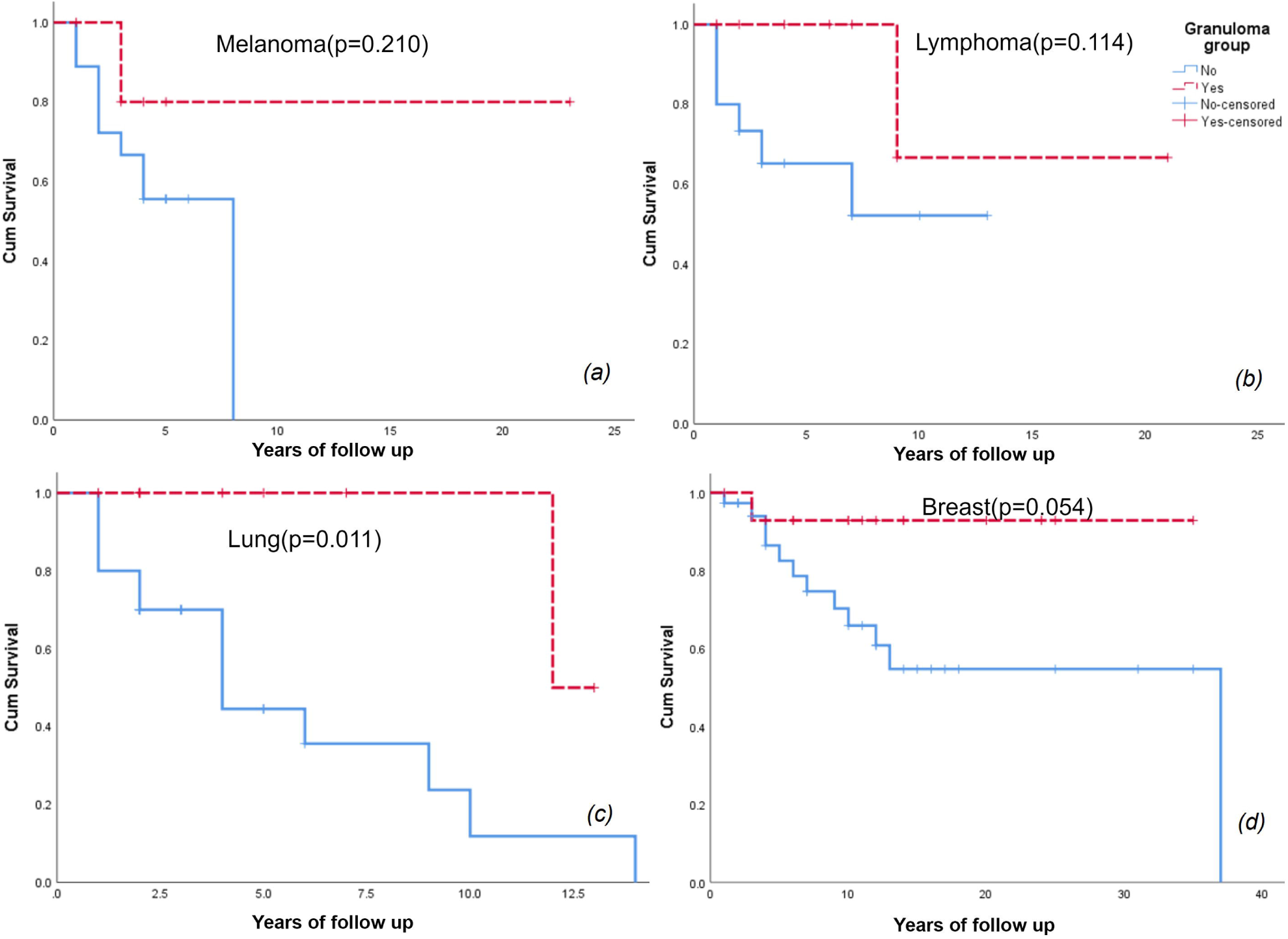
Logistic regression with stepwise elimination; p-value from Hosmer and Lemeshow goodness-of-fit test=0.497. The rhombus shape indicates the odds ratio (OR), and the vertical line indicates the confidence interval (CI). The arrow indicates that the 95% CI is outside the range shown.

### Survival analysis

The average number of years of follow-up for the cancer patients with and without sarcoidal granulomas was 7.58 years (SD=6.90) and 6.07 years (SD=6.49), respectively (p=0.284). Patients with both sarcoidal granulomas and cancer showed greater 2-year (OR=5.37 [1.35-21.4], p=0.017), 4-year (OR=4.49 [1.55-12.98], p=0.006), 6-year (OR=6.70 [2.42-18.61], p=<0.0001) and 10-year (OR=7.10 [2.80-18.00], p=<0.0001) survival times compared to those with cancer without the presence of sarcoidal granuloma (Table 2). However, there was no significant difference in survival between the two groups beyond 10 years (p=0.095).

**Table 2:** Comparison of cancer subjects with and without sarcoidal granuloma.

| Variable | Cancer only | Cancer and granuloma | p-value | OR (95% CI) |
| --- | --- | --- | --- | --- |
| Alive <2 years | 20(46.5%) | 14(82.4%) | 0.017 | 5.37 (1.35-21.4) |
| Alive <4 years | 40 (50%) | 23 (82.1%) | 0.006 | 4.49 (1.55-12.98) |
| Alive <6 years | 48 (51.1%) | 35 (87.5%) | <0.0001 | 6.70 (2.42- 18.61) |
| Alive <10 years | 57 (51.4%) | 45 (88.2%) | <0.0001 | 7.10 (2.80- 18.00) |
| Alive >10 years | 16 (69.6%) | 15 (93.8%) | 0.095 | 6.56 (0.72- 59.85) |
| Bladder Cancer | 7 (5.2%) | 3 (4.5%) | 0.819 | 0.85 (0.21- 3.40) |
| Breast Cancer | 37 (27.7%) | 15 (22.4%) | 0.426 | 0.76 (0.38-1.50) |
| Colon Cancer | 13 (6.5%) | 7 (5.2%) | 0.316 | 1.79 (0.58-5.54) |
| Lung Cancer | 30 (22.4%) | 10 (14.9%) | 0.215 | 0.61 (0.28-1.33) |
| Lymphoma | 15(11.2%) | 10 (14.9%) | 0.451 | 1.40 (0.59- 3.30) |
| Melanoma | 18 (13.4%) | 6 (9%) | 0.359 | 0.63 (0.24- 1.68) |
| Pancreatic Cancer | 4 (3%) | 3 (4.5%) | 0.589 | 1.52 (0.33- 7.01) |
| RCC | 8 (6%) | 3 (4.5%) | 0.662 | 0.74 (0.19- 2.88) |
| Sarcoma | 2 (1.5%) | 3 (4.5%) | 0.222 | 3.09 (0.50- 18.98) |
| Prostate Cancer | 6 (4.4%) | 8 (11.9%) | 0.059 | 2.89 (0.96- 8.71) |
| Cancer Stage 1, 2 and 3 | 53 (42.4%) | 35 (77.8%) | <0.0001 | 4.76(2.16-10.45) |
| Cancer Stage 4 | 72(57.6%) | 10 (22.2%) | <0.0001 | 0.21 (0.10-0.46) |
| Male | 53(39.6%) | 32 (47.8%) | 0.268 | 1.40 (0.77-2.52) |
| European-American | 56 (41.8%) | 23 (34.3%) | 0.308 | 0.73 (0.40-1.34) |
| Hispanic | 57(42.5%) | 17 (25.4%) | 0.019 | 0.46 (0.24- 0.88) |
| African American | 17 (12.7%) | 12 (17.9%) | 0.323 | 1.50 (0.67- 3.36) |
| Asian | 2(1.5%) | 12 (17.9%) | 0.001 | 14.40(3.12-66.48) |
| Surgery | 86(67.2%) | 39 (63.9%) | 0.659 | 0.87 (0.46-1.64) |
| Radiotherapy | 69 (53.9%) | 23(37.7%) | 0.038 | 0.52 (0.28-0.97) |
| Chemotherapy | 90 (70.3%) | 28 (45.9%) | 0.001 | 0.36 (0.19-0.67) |
| MAB | 55 (43.0%) | 12 (19.7%) | 0.002 | 0.33 (0.16- 0.67) |

OR=Odds ratio, CI=Confidence interval, RCC=Renal cells carcinoma, MAB=Monoclonal antibody therapy

Likely due to the decrease in stage IV cancer diagnoses, we observed a statistically significant difference in the treatment administered, with fewer subjects having sarcoidal granuloma being treated with chemotherapy (p=0.001), monoclonal antibody therapy (MAB) (p=0.002) and radiotherapy (p=0.038) compared to controls. There was a significant difference in the number of Asian and Latino subjects between the two groups, as the control group was obtained from the University of Miami database, which serves a predominantly Latino population with a relatively small Asian community.

Similarly, a multivariate Cox proportional hazards regression model adjusted for potentially confounding baseline demographic and clinical characteristics, including age, sex, ethnicity, cancer treatment and cancer stage, showed increased survival among subjects with sarcoidal granuloma and cancer compared to those with cancer alone (Figure 3).

**Figure 3:**
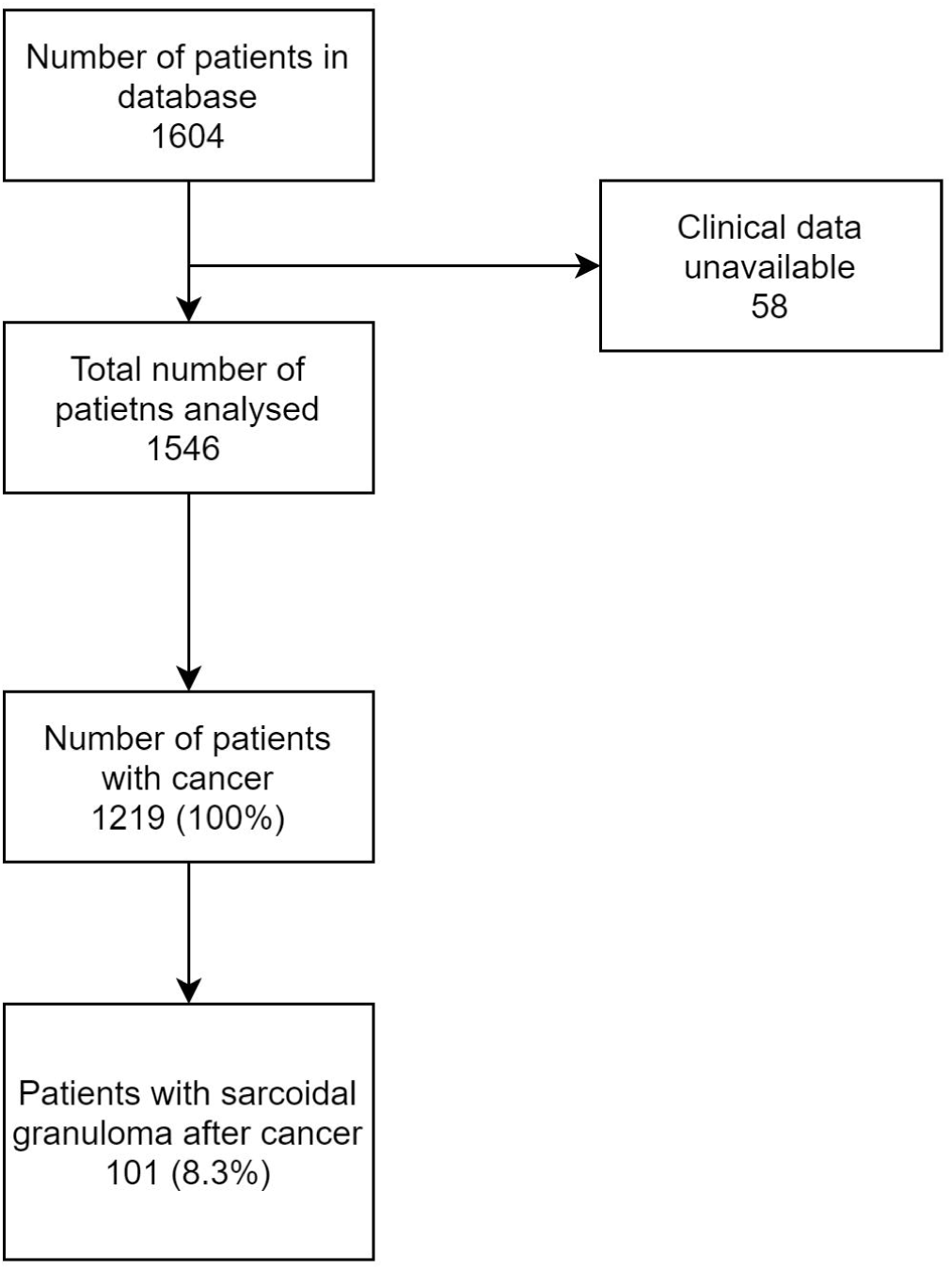
Multivariate Cox proportional hazard analysis of survival of cancer patients with and without sarcoidal granulomas.

Kaplan-Meier survival curves along with the log-rank test comparing cancer patients with and without sarcoidal granulomas for the most commonly occurring cancers (breast, lung, lymphoma and melanoma) in our dataset revealed only a significant survival advantage for those with lung cancer (p=0.011) (Figure 4), but not for melanoma (p=0.210), lymphoma (p=0.114) or breast cancer (p=0.054).

**Figure 4:**
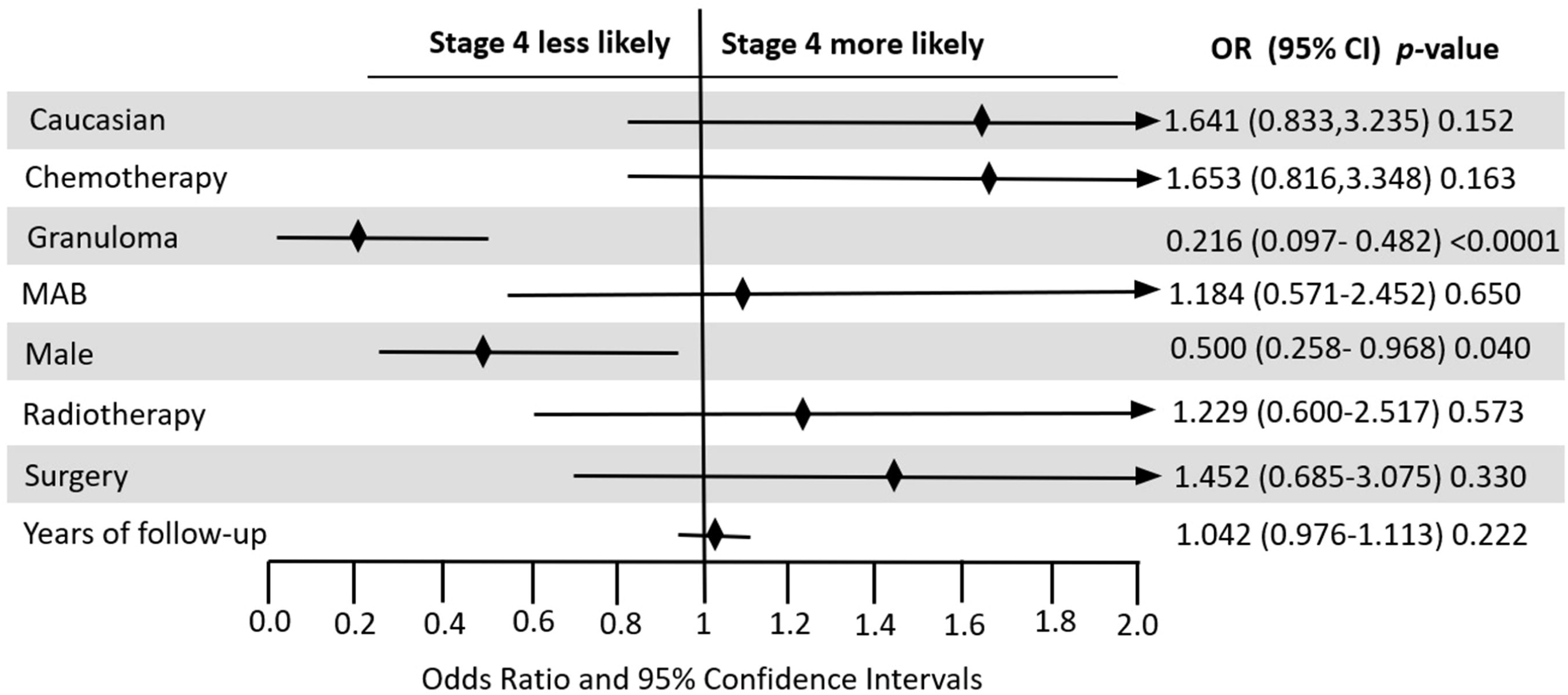
Kaplan-Meier survival analysis showing the survival curve for the most common cancers among controls and cases-Melanoma (a), Lymphoma (b), Lung cancer (c) and Breast cancer (d). The curves illustrate the proportion of subjects at each time point who were alive. The p-value was calculated by the log-rank test.

## Discussion

The current study found that 8.3% of cancer patients had sarcoidal granuloma in pathology samples. Patients with skin, breast, and prostate cancers and lymphoma were more likely to develop sarcoidal granuloma. Of most importance, this study found a decreased incidence of stage 4 cancer in patients with sarcoidal granulomas compared to matched controls and led to a significant survival advantage at 2, 4, 6 and 10 years.

Despite the wide range of cancers reported for individuals with sarcoidal granulomas, breast malignancy is the most commonly occurring cancer in most studies. A study by Keiss *et al*. investigated 64 patients with sarcoidal granuloma after cancer found 17% of the subjects had breast cancer[12]. Furthermore, Butt *et al*. reviewed 30 subjects with sarcoidal granuloma following cancer, of whom 33% had breast cancer[13]. A similar predominance of breast cancer has been recorded by several other authors[14–16], and this observation is consistent with our data.

The mean age of onset of sarcoidosis among our subjects was 64.5 years. This is in agreement with multiple studies describing the occurrence of sarcoidal granuloma in cancer patients is > 50 years instead of the typical ages of sarcoidosis patients who are between 20 and 39 years[7, 12, 15–17]. As suggested by Grados *et al*.[18] and Hunt *et al*.[14], the reason may be that cancer occurs more commonly in those of an older age. This discrepancy in age of diagnosis suggests that the malignancy may be providing the antigen that fuels the sarcoid-like granulomatous reaction.

Our findings also suggest a decreased incidence of stage 4 metastatic disease in patients with sarcoidal granulomas. Grados *et al*. in their analysis also showed that none of 12 subjects with sarcoidal granuloma presented with metastatic disease[18]. These findings may suggest a protective role for sarcoidal granulomas with regard to metastasis in cancer patients. This effect may be due to cancer cells being prevented from evading activated immune cells when there is a high tendency to develop granulomas. Further investigation of this speculation may lead to new therapeutic agents for cancer immunotherapy.

Overall, the prognostic value of sarcoidal granuloma in cancer requires further investigation. A recent study by Steinfort *et al*. among subjects with NSCLC indicated a significant cancer-free survival in those with sarcoidal granuloma *(n=8)* who had no recurrence of the disease compared to 44% of controls who had recurrence at a median interval of 11 months after surgery[7]. Similarly, in their study of survival among 19 patients with lung cancer and coexisting granulomatous inflammation, Dagaonkar *et al*. observed a 3-year survival rate of 21% compared to 6% in those without granulomas[19]. Regardless, several other studies failed to reveal any significant difference in prognosis in patients with sarcoidal granulomas[8, 20]. One major drawback of these studies is the relatively small sample sizes and univariate analysis of survival, which makes it challenging to predict prognosis accurately.

In our study, the overall survival for all patients with cancer and sarcoidal granuloma was significantly higher than that of the controls when analyzed using the Cox proportional hazards regression model. Significance was not reached in Kaplan-Meier analysis for survival for every individual cancer, except for lung cancer, which was probably due to the limited sample size; however, the trend for improved survival was apparent. The current data indicated a lower percentage of stage 4 disease in subjects with sarcoidal granuloma compared to controls, and this, in turn, might be the reason for the observed increase in survival.

Earlier studies have suggested chemotherapeutic agents as potentially inducing sarcoidal granulomas[21, 22]. We find this suggestion unlikely, as less than half of our subjects were treated with chemotherapy and not one chemotherapeutic agent was used in more than 12%.

One question that arises is the mechanism of granuloma formation. The immunology of sarcoidal reactions occurring in cancer patients remains undiscovered. Nonetheless, it has been suggested that these sarcoidal granulomas could be an immune reaction to cancer cells[13]. For this to be true, the antigen sustaining the granulomas must be derived from cancer cells. We suggest that vimentin may be a possible candidate antigen. Vimentin, a member of the intermediate filament family of proteins, has been identified in BAL samples from sarcoidosis subjects[23, 24], and the T-cells derived from these BAL samples have revealed identical T-cell receptor sequences. Interestingly, modeling of these sequences with HLA-DR3 protein sequences showed an ideal fit for a vimentin peptide[25]. In addition, macrophages, an essential part of sarcoid granulomas, have also been shown to secrete vimentin when activated[26]. The Kveim-Siltzbach agent, which was historically used in the diagnosis of sarcoidosis, contained vimentin, further implicating a role in sarcoidal granuloma formation[27].

Although vimentin is a major component of the cytoskeleton of mesenchymal cells, its expression is upregulated in various cancers of epithelial origin. Vimentin expression has also been reported in malignant lymphomas[28]. Its role in the metastatic cascade has also been well documented [29]. It is possible that vimentin from the primary tumor or metastatic cell triggers an immune response similar to sarcoidosis in genetically susceptible individuals, resulting in the formation of noncaseating granulomas. Currently, no studies assessing the role of vimentin in cancer-related sarcoid reactions have been conducted to corroborate this hypothesis.

Ideally, larger international prospective studies to analyze the incidence and determine the prognostic value of sarcoidal granulomas more accurately should be conducted. It is also essential to obtain a deeper understanding of the pathophysiology of cancer-related granulomas. Immune profiling of granulomas to assess the presence or absence of various receptors and markers, such as vimentin, maybe the first step. If these granulomas are indeed antitumor responses to metastasizing cells, studies to molecularly detect neoplastic cells within granulomas using techniques such as PCR can be performed[30].

### Limitations

Our study has all the limitations pertinent to retrospective studies. Despite being a relatively large study on this subject, we were unable to perform multivariate Cox survival analysis for every individual cancer due to the small subgroup sample size. One of our datasets consisted of melanoma patients only (140 subjects), which might be the reason for the higher number of subjects with a history of melanoma.

## Conclusion

The findings of our study suggest that cancer patients who develop sarcoidal granulomas have a survival advantage with lower rates of stage 4 disease. Thus, finding sarcoidal granulomas could serve as a prognostic biomarker. Although the exact mechanism of granuloma formation is unknown, they are likely to be an immune response to neoplastic components and if proven, could also serve as a therapeutic biomarker for immunotherapy. Larger case-control studies are required to confirm and further assess the increased survival reported herein and evaluate the presence of possible cancer-related biomarkers in these granulomas via molecular-based studies.

## Data Availability

The data that support the findings of this study are available from the corresponding author, upon reasonable request.

